# Hydroxyzine in Acute Hypertension: A Cohort Study

**DOI:** 10.1101/2024.09.30.24314673

**Authors:** Jakub Nożewski, Piotr Remiszewski, Kinga Filipek, Agata Pisklak, Aleksandra Abramczyk, Stanisław Pyzel, Daniel Śliż

## Abstract

Captopril, an ACE inhibitor, is widely used for acute hypertensive episodes, while hydroxyzine, an antihistamine with sedative and anxiolytic effects, is also employed in Poland despite limited research on its efficacy for this purpose. To our knowledge, no prior studies have specifically investigated the direct impact of hydroxyzine on hypertension, despite its common use in Polish medical practice.

A retrospective cohort analysis of 2144 patients who required emergency intervention for hypertension or had high blood pressure during other interventions. Effectiveness was based on blood pressure improvement post-intervention.

84.28% of interventions were performed by basic emergency teams, and 15.72% by specialist teams. Captopril and hydroxyzine were the most commonly used medications. Blood pressure improved in 36.61% of patients, with no improvement in 41.47%. Hydroxyzine was more frequently administered by paramedic teams, while captopril was favoured by specialist teams. There were no statistically significant differences in captopril use across age groups, with approximately 80% receiving it. Hydroxyzine use did not vary significantly by age, though its overall use was low. Hydroxyzine showed a non-significant trend towards greater blood pressure reduction compared to captopril.

Captopril is the primary treatment for acute hypertension in emergency settings, but hydroxyzine may have potential benefits, especially in patients with anxiety-related hypertension. However, due to the lack of significant evidence and current guidelines advising against hydroxyzine in elderly patients, further research is needed to establish protocols and optimize hypertension treatment strategies. Additionally, incorporating anxiolytics into guidelines and an official statement on their use would aid clinical practice.

## Introduction

According to the Polish Central Statistical Office, in 2022^1^, medical rescue teams made almost 3.1 million trips or departures to the emergency scene, of which 6.7% were children and adolescents under 18 years of age, and 48.5% were people aged 65 and over. The elderly accounted for almost half of those calling for the Emergency Medical Services Team. 1,592 Medical Rescue Teams were operating within the State Medical Rescue System in 2022, of which 1,271 were basic and 321 specialised. One very common reason for calling an Emergency Medical Response Team is a sudden increase in blood pressure.^2^

Hypertension is defined as office-measured systolic blood pressure (sBP) of ≥ 140 mmHg and/or diastolic blood pressure (DBP) of ≥ 90 mmHg. The World Health Organization (WHO) has identified hypertension as the leading cause of premature death worldwide. In 2015, the global prevalence of hypertension was estimated to be 1.13 billion.^3^ Among hypertensive patients calling the emergency medical team, we can distinguish two conditions: emergency and urgent. Emergency conditions in hypertension are defined as situations in which significantly elevated blood pressure (BP) values are accompanied by the development of acute organ complications, systolic blood pressure is generally over 180 mmHg, and diastolic blood pressure is over 110 mmHg.^4^ Patients report a range of symptoms, including headaches, visual disturbances, chest pain, shortness of breath, dizziness and neurological deficits. These patients may develop changes in the fundus of the eye, microangiopathy, disseminated intravascular coagulation (DIC), encephalopathy, acute heart failure, and acute impairment of renal function. The emergency is associated with the need to administer drugs intravenously.^5^ The drugs used in this case include β-blockers (esmolol, metoprolol, labetalol), fenoldopam, clevidipine, nicardipine, nitroglycerin, sodium nitroprusside, rapidly, clonidine, phentolamine, enalaprilat, among others. In an emergency, enalaprilat is the only angiotensin-converting enzyme inhibitor (ACEI) drug that can be used. The European Society of Cardiology guidelines do not recommend the use of captopril and hydroxyzine in hypertension-related emergencies.^6^ Hypertension urgency differs from emergency hypertension in the absence of acute organ complications. In these patients, hospitalization is not required, and blood pressure reduction can be carried out by oral route. In this case, the drug groups used are ACE inhibitors, ARBs or β-blockers. In an urgency, we can use captopril orally or sublingually.^7^ The guidelines do not provide any information on the use of hydroxyzine in treating acute hypertension.^1^

Angiotensin-converting enzyme inhibitors (ACEIs) represent a pivotal class of pharmaceuticals in the field of medicine. They are employed in the treatment of hypertension, heart failure, ischemic heart disease, and certain kidney diseases.^8^ Their mechanism of action involves the inhibition of the angiotensin-converting enzyme, which converts angiotensin I into angiotensin II. Angiotensin II raises blood pressure, mainly through vasoconstriction and by stimulating aldosterone secretion in the renin-angiotensin-aldosterone (RAA) system.^9^ Furthermore, the body increases the levels of angiotensin I, which in turn increases the release of nitrogen oxide, which has a vasodilatory effect on the blood vessels. Captopril belongs to the ACEI group and is one of the most commonly used drugs in this group for the treatment of hypertension, either as monotherapy or in combination with other drugs. It was the first drug from the ACEI group to be introduced to the pharmacological market; the drug was developed in 1975.^10^ When administered orally, it is absorbed in 75%.^11^ In Polish conditions, it is frequently used in sublingual or oral form for acute conditions associated with an increase in blood pressure. It has been demonstrated that the efficacy of the drug does not vary according to the route of administration, whether orally or sublingually.^12^ A less common drug administered to patients with sudden increases in blood pressure by emergency medical teams is hydroxyzine. Hydroxyzine is not typically used as a first-line treatment for sudden increase in blood pressure. It is primarily an antihistamine medication used to treat allergic reactions, itching, and anxiety. While it may have some sedative effects, it does not have significant antihypertensive properties. However, there are scenarios where anxiety or stress can contribute to elevated blood pressure readings. In such cases, if anxiety is a contributing factor to hypertension, hydroxyzine may be prescribed to help manage anxiety symptoms, potentially leading to a reduction in blood pressure.

## Methods

The study has been using retrospective cohort analysis. The provided data pertained to 2188 patients. We included data from 14 teams consisting of 2 paramedics and 3 teams consisting of a paramedic, a nurse, and a doctor, who were dispatched in 2019 to assist patients due to hypertension or other illnesses, but in the latter case, hypertension was also noted. We included the data from. The dosages for captopril were 25 mg and 12.5 mg. The data for this study was collected from 15 Basic Medical Rescue Teams (Podstawowe Zespoły Ratownictwa Medycznego) and 3 Specialist Teams (Zespoły Specjalistyczne). These teams operate under the Provincial Ambulance Station (Wojewódzka Stacja Pogotowia). For 57 individuals, the data were incomplete – these individuals did not have a registered PESEL number. Consequently, it was not possible to automatically determine the gender and age of these patients. Gender and age were established for 13 of them based on information entered in one of the columns, i.e., in the interview section. The remaining 44 patients without specified gender and age were preliminarily excluded from the analysis. Ultimately, the analysis covered 2144 patients. A significance level of 0.05 was adopted for the conducted tests.

Due to the left-skewed asymmetry in the age distribution and the presence of outliers when analysing age, non-parametric methods need to be used. The Mann-Whitney U test yielded U = 275008.500, where p < 0.001. These results suggest that there is a statistically significant difference in the median ages between the groups under investigation. In addition to p-values, we calculated Cohen’s d to quantify the effect size of the treatments. Cohen’s d is a measure of the strength of the treatment effect, providing insight into the practical significance of the findings beyond mere statistical significance.

The first group of variables are quantitative variables. These include the ones related to pulse, glucose level, systolic blood pressure, and diastolic blood pressure. Pulse and glucose were measured once, while blood pressure was measured up to three times, hence resulting in 6 variables related to blood pressure: systolic pressure1, systolic pressure2, systolic pressure3, diastolic pressure1, diastolic pressure2, diastolic pressure3.

The second group consists of qualitative variables. Among them is the type of ambulance. The basic team (Team P) was called for 1807 individuals (84.28%), while the specialised team (Team S) was called for the remaining 337 individuals (15.72%).

The study also employed CART decision tree analysis to ascertain the factors influencing whether patients were transferred to the hospital or not. The process involved dividing the dataset into 10 parts for 10-fold cross-validation. CART trees were built on each training set, considering variables such as systolic and diastolic improvements, gender, age, EMT type, pulse, glucose, improvement status, and reason for the call. The model yielded 10 trees, and their accuracy, sensitivity, and specificity were determined. Additionally, the study utilised ROC curve analysis to evaluate classification quality, determining the area under the curve (AUC) as a measure of model performance. The importance of predictors was assessed, revealing the most influential variables.

Chi-square tests were employed to assess the independence of variables, revealing a significant association (p < 0.001) between the reason for the call and hospital transfer. Further analyses examined the independence of hospital transfer from variables such as gender, EMT type, age, pulse, and blood pressure measurements, separately for calls related to hypertension and other illnesses.

## Results

We analyzed the therapeutic management applied by emergency medical teams called for intervention due to high blood pressure values or for other reasons, but high blood pressure values were noted during the intervention. This study aimed to analyze the effectiveness and appropriateness of pharmacological interventions used in such cases. We aimed to analyze the effectiveness of emergency medical teams in managing hypertension emergencies and explore possibilities for optimizing patient care in similar circumstances.

The study was conducted on a sample of 2188 patients, 2144 of whom were included in the study. The study included 1597 women and 547 men. It noted that emergency services were called approximately three times more often for women than for men, which may suggest gender differences in health issues or perception of symptoms. Female participants’ ages range from 19 to 99 years, with a mean age of 74.63 years and a standard deviation of 12.36 years. The median age for female participants was 77 years, reflecting a slight skew towards older ages within the cohort. Male participants had an age range of 20 to 97 years, with a mean age of 64.88 years and a standard deviation of 15.77 years. The median age for male participants was 67 years.

Quantitative variables such as blood pressure values from three measurements, heart rate values and glucose levels were analysed—table A.

**Table A.** Summary statistics of quantitative variables. N - The number of patients for whom a particular measurement was taken. Min – Max - Minimum and maximum values

| Variables | N | Average ± standard deviation | Min – Max | Median |
| --- | --- | --- | --- | --- |
| Heart rate | 2125 | 85,73 ± 16,31 | 40 – 180 | 82 |
| Glucose | 1201 | 138,86 ± 50,83 | 32 – 600 | 126 |
| Systolic blood pressure 1. | 2133 | 191,53 ± 22,71 | 107 – 300 | 190 |
| Systolic blood pressure 2. | 880 | 161,93 ± 22,18 | 110 – 250 | 160 |
| Systolic blood pressure 3. | 134 | 163,84 ± 25,41 | 120 – 260 | 160 |
| Diastolic blood pressure 1. | 2128 | 97,96 ± 13,10 | 60 – 170 | 100 |
| Diastolic blood pressure 2. | 879 | 87,29 ± 11,64 | 50 – 165 | 90 |
| Diastolic blood pressure 3. | 134 | 87,17 ± 11,76 | 60 – 140 | 85 |

We analysed the effectiveness of an intervention in which captopril and hydroxyzine were administered. 1710 (79,76%) patients received captopril. Hydroxyzine was administered to 152 people (7.09%). Both drugs were given simultaneously to 128 people (5.97%). The analysis showed that the applied treatment improved blood pressure parameters in 36.61% of patients, while a lack of improvement was observed in 41.47% of individuals.

The majority of patients, 84.28%, received intervention from the basic team B-type (paramedics only), while the remaining 15.72% were serviced by a specialist team S-type (paramedics and a doctor). Significant differences in the treatment administered were observed based on the type of emergency medical services team and the reason for the emergency call. There are no statistically significant differences in the use of Hydroxyzine depending on the type of emergency medical team. Captopril, on the other hand, is more often recommended by B-type teams (p = 0,021). Furthermore, both Captopril and Hydroxyzine were more frequently administered to patients with emergency calls due to hypertension Table B.

**Table B.** Summary of the reason for the call and the treatment used.

| Drug | The reason for the call |  | p-value (statistics) |
| --- | --- | --- | --- |
|  | Another reason<br>(n=1018) | High blood<br>pressure (n=1126) |  |
| Captopril | 771 (75,74%) | 939 (83,39%) | <0,001 (18,938) |
| Hydroxyzine | 48 (4,72%) | 104 (9,24%) | <0,001 (15,912) |

No statistically significant differences were observed in the administration of Captopril by age group. In each of the study groups, the percentage of patients who received Captopril was about 80%. Moreover, there are also no statistically significant differences in the frequency of Hydroxyzine administration among patient groups by age. However, the percentage of patients who were given this drug is not high. Table C.

**Table C.** Use of Captopril and Hydroxyzine in groups distinguished by patient age.

| Drug | <65 years old<br>(n=529) | 65-79 years old<br>(n=894) | >80 years old<br>(n=721) | p-value (statistics) |
| --- | --- | --- | --- | --- |
| Captopril<br>n (%) | 415 (78,4%) | 725 (81,1%) | 570 (79,1%) | 0,412 (1,772) |
| Hydroxyzine<br>n (%) | 31 (5,9%) | 76 (8,5%) | 45 (6,2%) | 0,095 (4,706) |

The decision to transfer patients to the hospital after emergency services intervention was significantly different depending on the reason for the emergency call. When the call was due to hypertension, 36.23% of patients were transferred to the hospital, while 55.80% were transferred when the call was for another illness (p < 0.001). Results indicated no influence of EMT type on hospital transfer, whether the call was for hypertension (p = 0.799) or another disease (p = 0.795). In cases where the intervention was due to hypertension, gender did not influence the decision to transfer to the hospital (p = 1). However, for calls related to other diseases, male patients were statistically significantly more likely to be transferred to the hospital (p = 0.027).

Regardless of whether the Emergency Medical Service team was called because of hypertension (p = 0.867) or another disease (p = 0.853), the patient’s age does not determine the transfer to the hospital. measurements were observed, indicating potential clinical relevance. Table D.

**Table D.** Statistics of quantitative characteristics of patients in the groups of transferred and non-transferred patients, divided into patients to whom the Emergency Medical Service was called for high blood pressure and another reason. n- number of patients

| The reason for the call | Factors | Transfer to the hospital |  | Student's t-test |  |  |
| --- | --- | --- | --- | --- | --- | --- |
|  |  | No | Yes | p-value | statistics | d Cohen |
| Another reason | Age | **n=450** | **n=568** | 0,853 | 0,185 | - |
| | | 70,1 ±16,17 | 69,9 ±14,55 | | | |
|  | Heart rate | **n=443** | **n=561** | <0,001 | -3,560 | -0,217 |
| | | 85,75 ±13,50 | 89,52 ±19,94 | | | |
|  | Systolic blood pressure 1. | **n=449** | **n=566** | <0,001 | -9,367 | -0,577 |
| | | 182,48 ±19,29 | 195,23 ±24,07 | | | |
|  | Systolic blood pressure 2. | **n=216** | **n=107** | <0,001 | -7,871 | -1,106 |
| | | 153,02 ±15,39 | 175,02 ±26,81 | | | |
|  | Systolic blood pressure 3. | **n=19** | **n=10** | 0,012 | -3,047 | -1,542 |
| | | 152,79 ±11,74 | 186,00 ±33,40 | | | |
|  |  | **n=449** | **n=567** | <0,001 | -5,470 | -0,338 |
|  | Diastolic blood pressure 1. | 95,94 ±11,62 | 100,35 ±14,03 |  |  |  |
|  | Diastolic blood pressure 2. | **n=217** | **n=106** | <0,001 | -6,344 | -0,883 |
|  |  | 84,52 ±9,47 | 94,96 ±15,60 |  |  |  |
|  | Diastolic blood pressure 3. | **n=19** | **n=10** | 0,056 | -1,999 | -1,999 |
|  |  | 84,84 ±8,56 | 95,00 ±19,00 |  |  |  |
| High blood pressure | Age | **n=718** | **n=408** | 0,867 | 0,167 | - |
|  |  | 74,1 ±12,16 | 74,2 ±12,67 |  |  |  |
|  | Heart rate | **n=711** | **n=407** | 0,020 | -2,336 | -0,160 |
|  |  | 83,17 ±12,20 | 85,48 ±17,74 |  |  |  |
|  | Systolic blood pressure 1. | **n=710** | **n=408** | <0,001 | -13,528 | -0,853 |
|  |  | 186,86 ±20,26 | 204,50 ±21,38 |  |  |  |
|  | Systolic blood pressure 2. | **n=450** | **n=107** | <0,001 | -11,071 | -1,645 |
|  |  | 157,03 ±15,67 | 187,42 ±27,35 |  |  |  |
|  | Systolic blood pressure 3. | **n=80** | **n=25** | <0,001 | -7,439 | -2,407 |
|  |  | 153,64 ±13,20 | 196,00 ±27,50 |  |  |  |
|  | Diastolic blood pressure 1. | **n=705** | **n=407** | <0,001 | -8,893 | -0,580 |
|  |  | 94,90 ±11,67 | 102,15 ±13,88 |  |  |  |
|  | Diastolic blood pressure 2. | **n=449** | **n=107** | <0,001 | -7,845 | -1,090 |
|  |  | 84,81 ± 8,93 | 95,74 ±13,74 |  |  |  |
|  | Diastolic blood pressure 3. | **n=80** | **n=25** | <0,001 | -5,203 | -1,446 |
|  |  | 83,55±8,52 | 97,40±12,43 |  |  |  |

This analysis is presented as a Classification and Regression Tree (CART) model for predicting hospital admission outcomes. The model classifies whether a patient will be admitted to the hospital (“YES”) or not (“NO”) based on various medical and demographic variables. The root node starts with 2148 cases, split into 54.6% “NO” and 45.4% “YES,” with an overall success rate of 0.180. The CART model identifies significant variables such as physical improvement, pulse rate, age, reason for EMS call, and improvement status, highlighting their roles in predicting hospital admissions. This model provides a structured approach to understanding the factors affecting patient outcomes, with potential applications in improving triage and patient management protocols.

The root node splits based on “physical improvement” at a threshold of 7.5. Patients with a value of 7.5 or less are further split by “pulse rate” at 113.5, while those above 7.5 are split by the “reason for EMS call,” specifically “other disease” and “hypertension.” For patients with a pulse rate of 113.5 or less, the model further splits based on “age” at 57.5. For those with a pulse rate above 113.5, the split is based on “improvement?” categorized as “discharge” or “NO, YES.” Each node demonstrates varying success rates, indicating the model’s ability to distinguish between factors influencing hospital admissions. For example, patients with “physical improvement” scores above 7.5 and called EMS for “hypertension” have a higher admission rate (54.9%). Conversely, older patients (above 57.5) with lower pulse rates are less likely to be admitted (31.7%).

To determine the factors influencing hospital admissions, the CART decision tree was constructed using a dataset divided into ten parts for ten-fold cross-validation. The tree was built with 90% of the observations, using variables such as the difference in systolic blood pressure, improvement in diastolic pressure, gender, age, type of EMS (P – S), heart rate, glucose level, improvement status (YES-NO-empty), and reason for EMS call (hypertension – another disease). The resulting ten trees provided values for accuracy, sensitivity, and specificity, with average values presented in Table 4.4: specificity 58.69%, sensitivity 71.87%, and accuracy 64.43%. The model’s accuracy indicates that it correctly predicts hospital admissions for nearly 65% of patients. Sensitivity shows that the tree accurately identifies patients who were admitted (approximately 72%), while specificity indicates that the model correctly identifies patients who were not admitted (about 60%).

In Figure A, two key decision thresholds are marked with orange points. These points have coordinates (0.41, 0.72), which are derived from default cutoff probability values. These coordinates represent the sensitivity (true positive rate) and 1-specificity (false positive rate) at these thresholds. The figure also includes a black point with coordinates (0.54, 0.89), indicating the optimal cutoff point determined by Youden’s index. Youden’s index maximizes the sum of sensitivity and specificity, providing the best balance between true positive and true negative rates. This point is significant because it helps identify the probability threshold where the model’s performance is most effective. At this threshold, the model correctly classifies 89% of the patients who need to be admitted and correctly identifies 54% of the patients who do not need to be admitted. The ROC (Receiver Operating Characteristic) curve associated with these points helps visualise the trade-off between sensitivity and specificity for different threshold values. The AUC (Area Under the Curve) of 0.722 indicates that the model has good discriminative ability, effectively distinguishing between patients who need hospital admission and those who do not.

Figure A illustrates the importance of predictors in the model, with variables like the difference in systolic blood pressure, diastolic improvement, improvement status, reason for EMS call, pulse rate, age, gender, and EMS type being significant. The decision points on the tree (Figure 4.2) highlight key thresholds, such as a physical improvement score of 7.5 and a pulse rate of 113.5, which are critical in the classification process. The ROC curve for the model shows an AUC of 0.722, indicating good classifier performance.

**Figure A.**
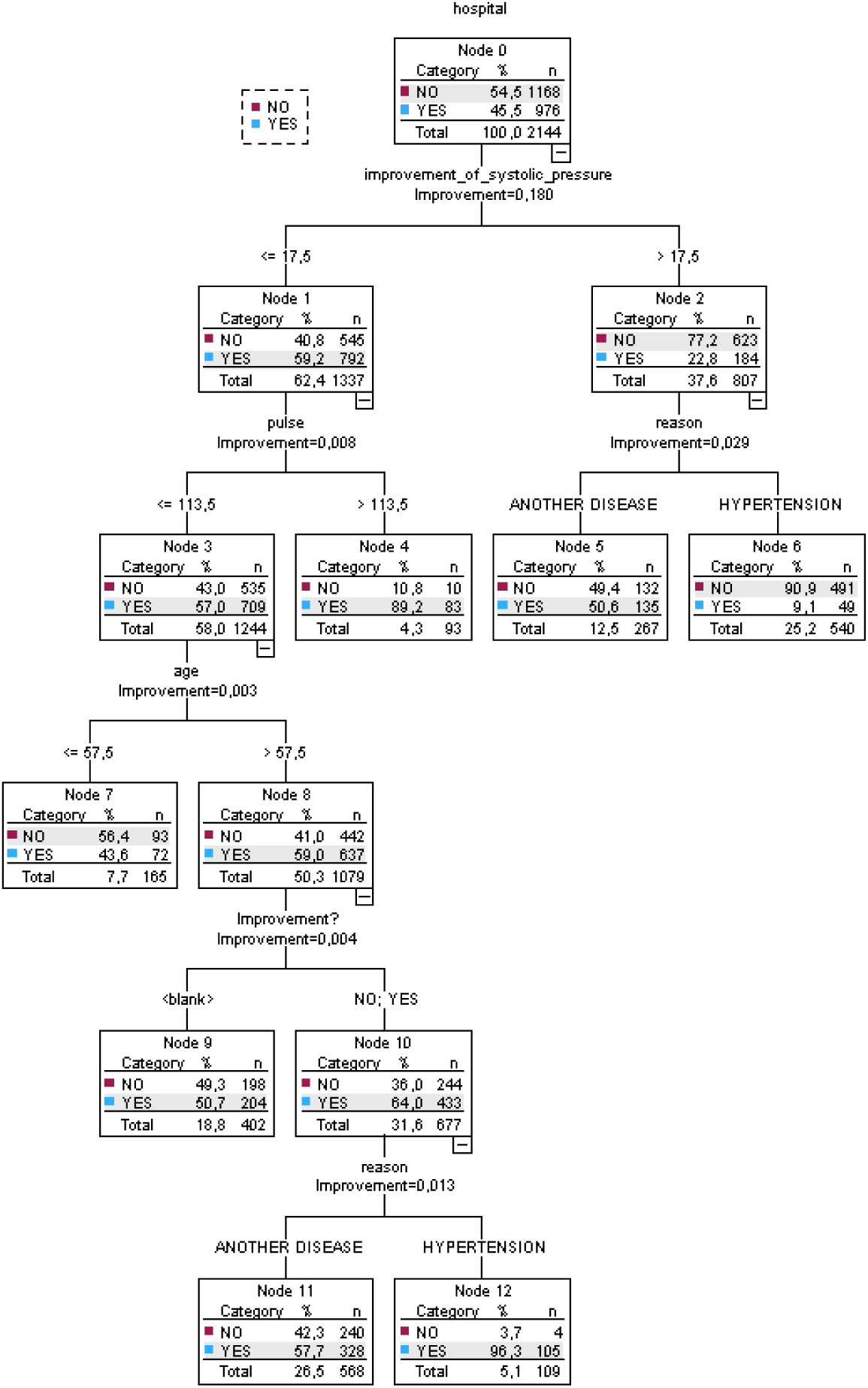
The effectiveness of the intervention was determined by physician assessment. It was found that in most cases 51,72%, administration of captopril did not result in improvement (p = 0,015). Conversely, in cases where hydroxyzine was administered, the percentage of patients not showing improvement was significantly lower at 42,98% (p = 0,025).

The improvement, defined as a reduction in blood pressure, was determined by the difference in values between the first and third blood pressure measurements. Reduction in blood pressure was observed regardless of whether pharmacotherapy was administered or not. Table 2.1.-2.4

**Table E.1.** Statistics for systolic blood pressure according to Captopril administration.

| Captopril | Systolic Blood Pressure 1. | Systolic Blood Pressure 3. | Student's t-test |  |
| --- | --- | --- | --- | --- |
|  |  |  | p-value | statistics |
| Not administered | 220,00 ± 29,28 | 165,63 ± 40,66 | 0,006 | 3,939 |
| Administered | 203,21 ± 19,83 | 163,72 ± 24,37 | <0,001 | 16,022 |

**Table E.2.** Statistics for diastolic blood pressure according to Captopril administration.

| Captopril | Diastolic Blood Pressure 1. | Diastolic Blood Pressure 3. | Student's t-test |  |
| --- | --- | --- | --- | --- |
|  |  |  | p-value | statistics |
| Not administered | 103,75 ± 28,25 | 93,13 ± 21,54 | 0,097 | 1,916 |
| Administered | 100,55 ± 13,61 | 86,72 ± 10,94 | <0,001 | 11,375 |

**Table E.3.** Statistics for systolic blood pressure according to Hydroxyzine administration.

| Hydroxyzine | Systolic Blood Pressure 1. | Systolic Blood Pressure 3. | Student's t-test |  |
| --- | --- | --- | --- | --- |
|  |  |  | p-value | statistics |
| Not administered | 204,73 ± 20,93 | 164,13 ± 26,22 | <0,001 | 15,255 |
| Administered | 201,05 ± 19,90 | 162,05 ± 20,25 | <0,001 | 5,873 |

**Table E.4.** Statistics for diastolic pressure according to Hydroxyzine administration.

| Hydroxyzine | Diastolic Blood Pressure 1. | Diastolic Blood Pressure 3. | Student's t-test |  |
| --- | --- | --- | --- | --- |
|  |  |  | p-value | statistics |
| Not administered | 100,21 ± 15,52 | 87,29 ± 11,63 | <0,001 | 9,943 |
| Administered | 103,95 ± 8,43 | 86,05 ± 13,18 | <0,001 | 6,470 |

We verified whether the administration of drugs resulted in higher values of pressure drops comparing when the drug was not administered. The administration of captopril had no statistically significant effect on the mean decrease in systolic (p = 0.153) or diastolic (p = 0.519) blood pressure. Similar conclusions were drawn for hydroxyzine concerning systolic (p = 0.822) and diastolic (p = 0.141) blood pressure. However, it should be noted that this inference may not be valid due to the small number of patients (134) whose blood pressure was measured three times, as well as the significant difference in the number of patients who received and did not receive the drugs.

## Discussion

The results indicate that captopril impacts blood pressure reduction in acute settings. Among the 2144 patients analysed, 36.61% showed improvement in blood pressure parameters post-administration. However, mean values of both systolic and diastolic blood pressure decreased regardless of whether which drug was administered to patients. In addition, the use of pharmacotherapy did not result in higher values of blood pressure drop, compared to the group that did not receive any medication. In our study, we observed that hydroxyzine, despite its anxiolytic properties, showed only a non-significant trend towards greater blood pressure reduction compared to captopril. This contrasts with findings from a related study on midazolam^13^, which demonstrated statistically significant reductions in systolic (P = 0.024), diastolic (P = 0.001), and mean blood pressure (P = 0.009) following treatment. In that study, the combination of midazolam and captopril resulted in the greatest BP reductions, with systolic, diastolic, and mean BP reductions of 23.5% (P = 0.047), 17.4% (P = 0.021), and 20.5% (P = 0.031), respectively. These results suggest that anxiolytics like midazolam may offer a more pronounced and rapid effect in lowering blood pressure than hydroxyzine.

Given midazolam’s effectiveness and faster onset in reducing BP, it raises important considerations about the role of anxiolytics in acute hypertension management. While hydroxyzine’s use in our cohort may be beneficial in anxiety-induced hypertension, the evidence for midazolam’s superior efficacy indicates that it could be a more appropriate choice in emergency settings, especially when quick BP reduction is critical.

Additionally, a study by Al-Mahmood et al. (2021) investigated the long-term use of angiotensin-converting enzyme (ACE) inhibitors, specifically lisinopril, and its impact on adrenergic receptor response in hypertensive patients. This research demonstrated that prolonged treatment with lisinopril significantly altered adrenergic receptor responses, which could enhance the management of hypertension. The findings emphasised the role of ACE inhibitors in modulating cardiovascular responses and improving outcomes for hypertensive patients, suggesting that these treatments lead to better blood pressure control and reduced risk of hypertensive crises. Consequently, the study concluded that ACE inhibitors like lisinopril are beneficial for the long-term management of hypertension, particularly by mitigating adrenergic overactivity associated with hypertension and its complications.

Moreover, Lin et al. (2021)^14^ explored the efficacy of the small molecule compound nerolidol against hypertension-induced cardiac hypertrophy in spontaneously hypertensive rats. The study showed that nerolidol administration significantly attenuated Angiotensin-II induced hypertrophic markers in cardiomyoblasts and reduced blood pressure in hypertensive rats. The beneficial effects of nerolidol were attributed to its ability to modulate the Mel-18-IGF-IIR signalling pathway, suggesting its potential as a therapeutic agent in managing hypertension-related cardiac complications.

Interestingly, the study found no statistically significant difference in captopril administration across different age groups, with about 80% of patients in each age category receiving the drug. However, considering the updated Beers criteria and other geriatric pharmacotherapy guidelines, caution is advised when administering captopril to elderly patients, particularly those over 80, due to the increased risk of adverse effects.

The study observed gender differences in the frequency of emergency calls and subsequent hospital transfers. Women were more likely to call for emergency services, but men were more often transferred to the hospital. This could suggest that men may present with more severe symptoms or that there are gender-based differences in symptom perception and reporting.

However, the gender of the patient did not influence the decision to transfer when the call was due to hypertension alone. Interestingly, a study by Todorov et al. (2021)^15^ investigated gender differences in intensive care unit treatment of patients, suggesting women were less likely to receive intensive care unit treatment regardless of disease severity. Gender differences were also noted in the prescription of antibiotics, Schröder et al. (2016)^16^ women received a significantly higher number of antibiotic prescriptions than men. It needs future investigation to provide equal health care regardless of the patient’s gender. The decision to transfer patients to the hospital was significantly influenced by the reason for the emergency call. When hypertension was the primary concern, 36.23% of patients were transferred, compared to 55.80% for other illnesses. This discrepancy highlights the need for further investigation into the criteria used for hospital transfers and suggests potential biases or gaps in the decision-making process of emergency medical teams.

Hydroxyzine is primarily indicated for anxiety disorders. Despite not being registered for the treatment of hypertension, it is sometimes used by emergency medical services teams in cases of emergency hypertension. This practice raises questions about the correctness of such interventions. Some research indicates that the intake of anxiolytics (e.g. diazepam, alprazolam) during blood pressure elevations can have beneficial effects, suggesting it might reduce unnecessary visits to the emergency room Tandeter et al. (2016)^17^. In our study hydroxyzine was administered to 152 patients, 68% of these administrations occurred when the primary reason for the call was high blood pressure. It is possible that the administration of hydroxyzine was motivated by reasons other than high blood pressure values, such as comorbid anxiety. Interestingly, most patients showed improvement and a drop in blood pressure after Hydroxyzine was administered. However, this reduction in blood pressure was observed regardless of whether pharmacotherapy was administered. This suggests the necessity for careful administration of drugs, as it exposes patients to potential adverse effects.

Based on the discussed studies, it is evident that both immediate and long-term management strategies are crucial for handling hypertensive emergencies and chronic hypertension. Midazolam offers rapid relief and is particularly useful in acute settings due to its fast action and dual role in reducing blood pressure and alleviating stress. ACE inhibitors like lisinopril provide sustained benefits by modulating adrenergic responses, thereby offering a preventive approach to managing hypertension. Nerolidol’s potential to reduce hypertrophic effects through specific signalling pathways further enriches the therapeutic landscape. The combined use of rapid-acting agents like midazolam, sustained therapies such as ACE inhibitors, and novel compounds like nerolidol provides a robust framework for effective hypertension management.

### Hypertension management guidelines

Guidelines for treating high blood pressure vary from country to country. One article collected 48 guidelines from around the world. Analysis of these guidelines showed that, in most cases, the guidelines agreed on the definition, severity and target blood pressure recommendations for hypertension, but the main differences were in the pharmacological strategies for treating hypertension. The most commonly recommended antihypertensive drug classes were angiotensin-converting enzyme inhibitors (ACEi), angiotensin receptor blockers (ARBs), beta-blockers, calcium channel blockers (CCBs) and diuretics. Of the 47 guidelines that recommended ACEi or ARBs, 68% did not recommend either. The remaining 32% of guidelines recommended ACEi over ARBs in patients with uncomplicated hypertension.^18^ A separate group of patients are geriatric patients, i.e. patients over the age of 65. In 1991, Mark Beers and co-workers developed the criteria, the essence of which is the identification of drugs that have a negative impact on the health of older people. In 2023, the American Geriatric Society (AGS) published another update of the Beers Criteria, a list of drugs that may be inappropriate for older patients. These drugs include ACEIs and hydroxyzine. In our study, we compared the management of hypertension in geriatric patients according to the European Hypertension Treatment Guidelines with the Beers Criteria. (Tab.)

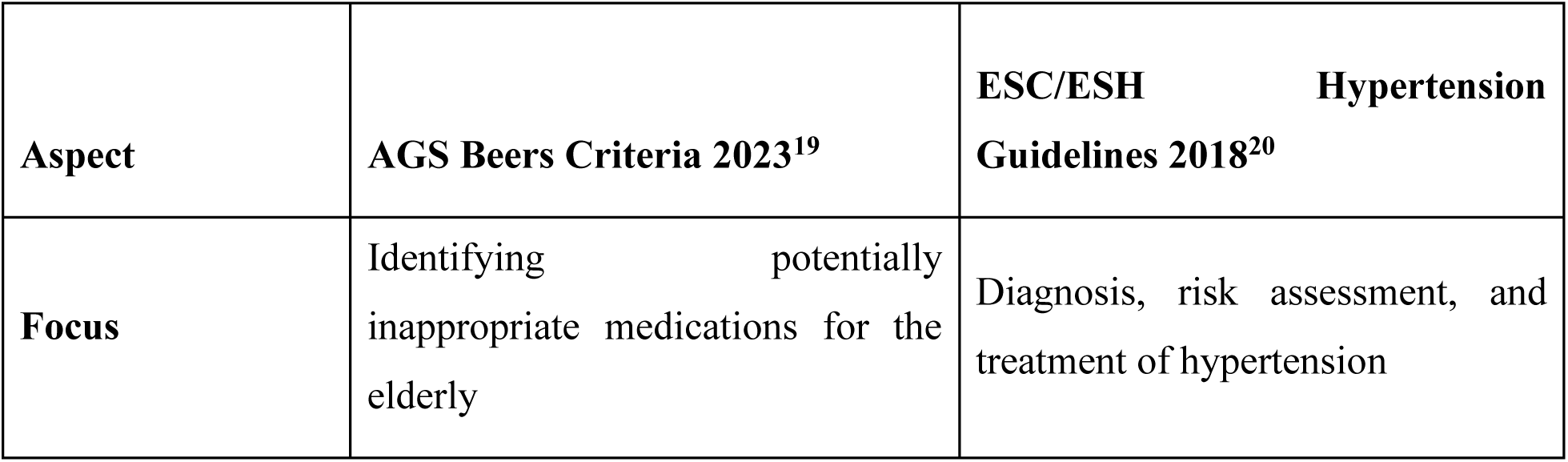
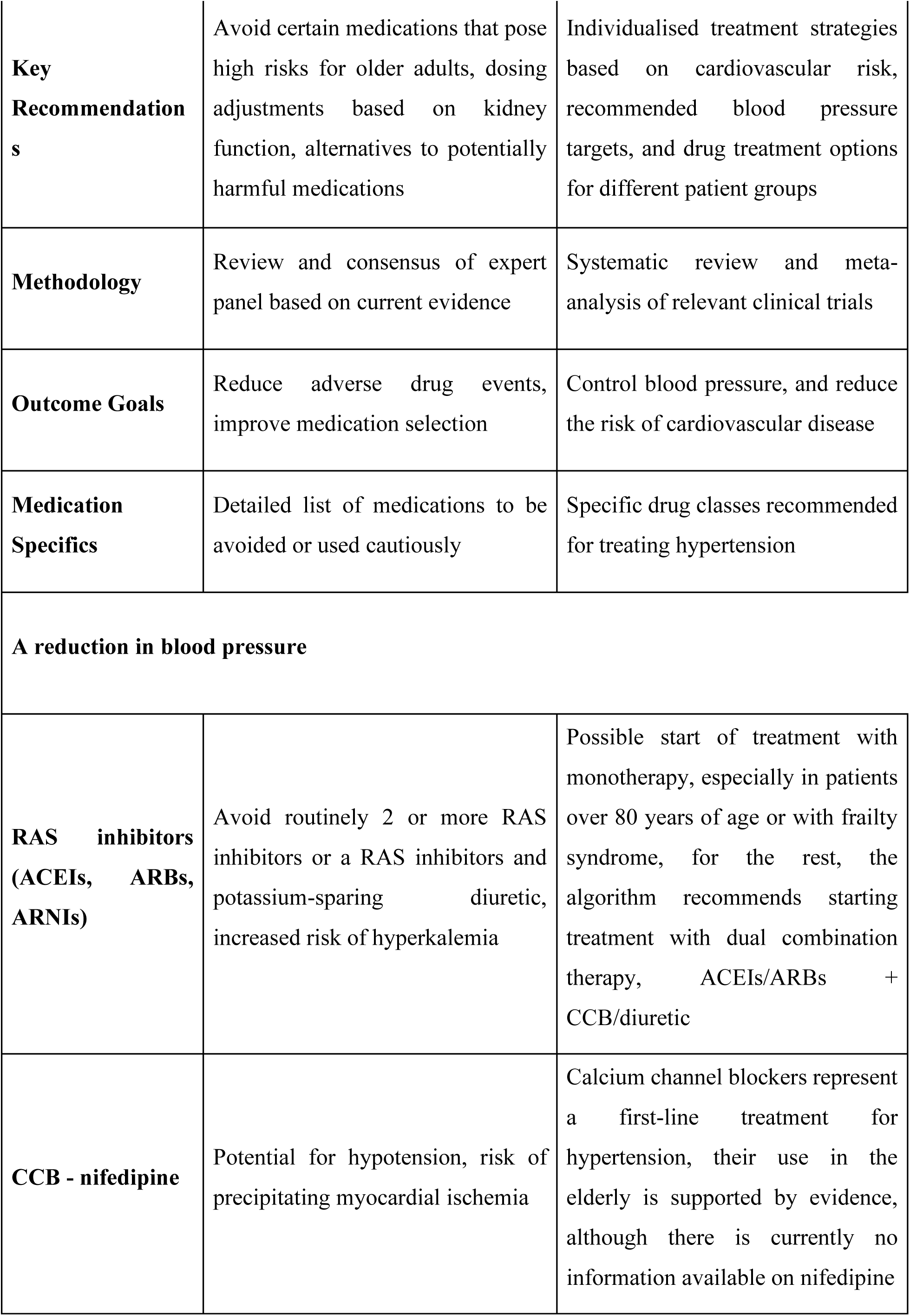
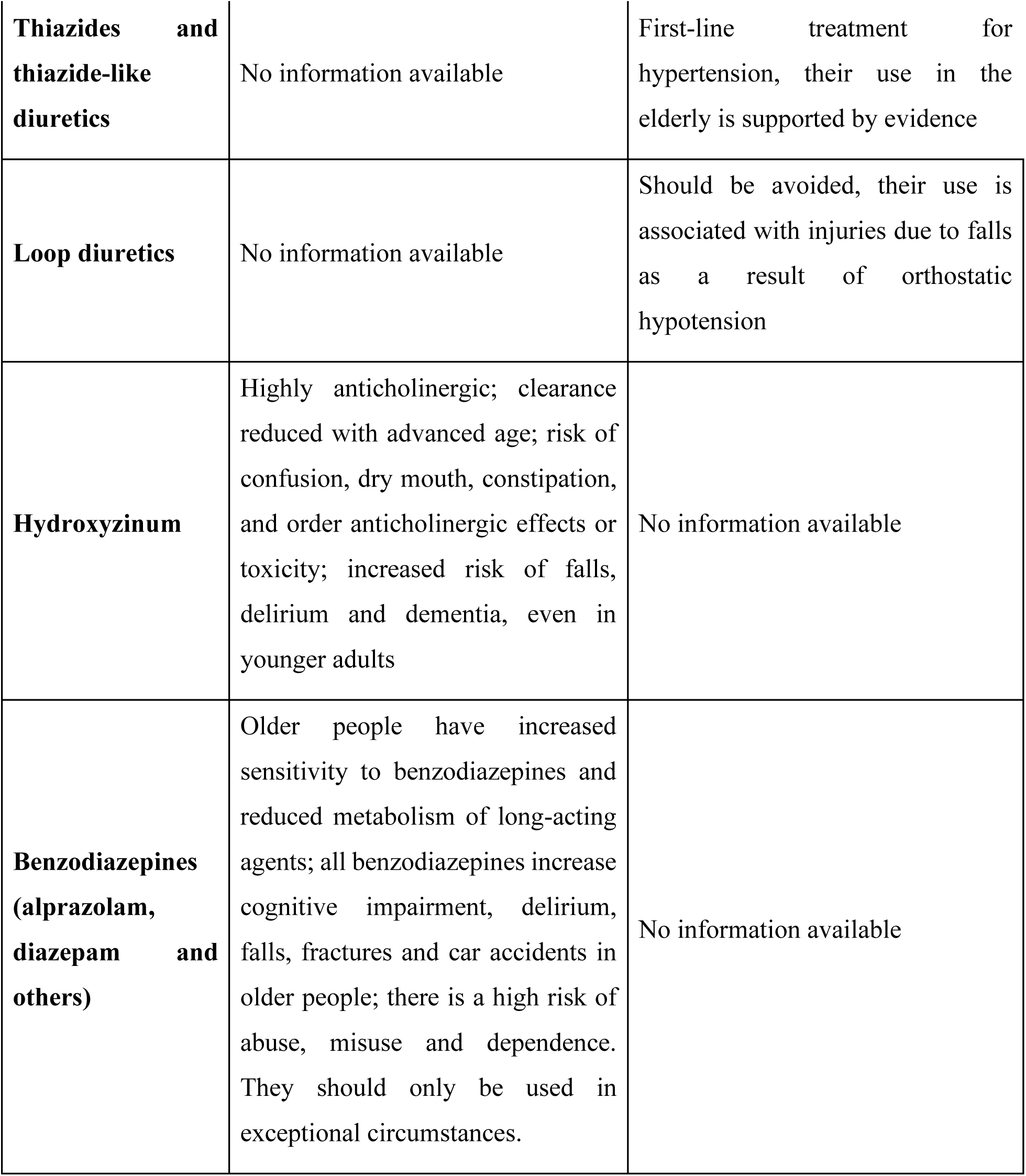
Tab. Comparison of AGS Beers Criteria 2023 with ESC/ESH Hypertension Guidelines 2018

| **Aspect** | **AGS Beers Criteria 2023<sup>19</sup>** | **ESC/ESH Hypertension Guidelines 2018<sup>20</sup>** |
| --- | --- | --- |
| **Focus** | Identifying potentially inappropriate medications for the elderly | Diagnosis, risk assessment, and treatment of hypertension |
| **Key Recommendations** | Avoid certain medications that pose high risks for older adults, dosing adjustments based on kidney function, alternatives to potentially harmful medications | Individualised treatment strategies based on cardiovascular risk, recommended blood pressure targets, and drug treatment options for different patient groups |
| **Methodology** | Review and consensus of expert panel based on current evidence | Systematic review and meta-analysis of relevant clinical trials |
| **Outcome Goals** | Reduce adverse drug events, improve medication selection | Control blood pressure, and reduce the risk of cardiovascular disease |
| **Medication Specifics** | Detailed list of medications to be avoided or used cautiously | Specific drug classes recommended for treating hypertension |
| **A reduction in blood pressure** |  |  |
| **RAS inhibitors (ACEIs, ARBs, ARNIs)** | Avoid routinely 2 or more RAS inhibitors or a RAS inhibitors and potassium-sparing diuretic, increased risk of hyperkalemia | Possible start of treatment with monotherapy, especially in patients over 80 years of age or with frailty syndrome, for the rest, the algorithm recommends starting treatment with dual combination therapy, ACEIs/ARBs + CCB/diuretic |
| **CCB - nifedipine** | Potential for hypotension, risk of precipitating myocardial ischemia | Calcium channel blockers represent a first-line treatment for hypertension, their use in the elderly is supported by evidence, although there is currently no information available on nifedipine |
| **Thiazides and thiazide-like diuretics** | No information available | First-line treatment for hypertension, their use in the elderly is supported by evidence |
| **Loop diuretics** | No information available | Should be avoided, their use is associated with injuries due to falls as a result of orthostatic hypotension |
| **Hydroxyzinum** | Highly anticholinergic; clearance reduced with advanced age; risk of confusion, dry mouth, constipation, and other anticholinergic effects or toxicity; increased risk of falls, delirium and dementia, even in younger adults | No information available |
| **Benzodiazepines (alprazolam, diazepam and others)** | Older people have increased sensitivity to benzodiazepines and reduced metabolism of long-acting agents; all benzodiazepines increase cognitive impairment, delirium, falls, fractures and car accidents in older people; there is a high risk of abuse, misuse and dependence. They should only be used in exceptional circumstances. | No information available |

CCB- calcium channel blockers

CCB- calcium channel blockers
|  |  |  |
| --- | --- | --- |
| **Anxiolytic** | **Effectiveness in reducing blood pressure** | **Pressure-reducing mechanism** |
| **Hydroxyzinum** | Hydroxyzine is primarily used to treat anxiety disorders, but it is sometimes employed off-label for emergency hypertension management by emergency medical services. Its effectiveness in reducing blood pressure, however, is not well-supported by statistical evidence. The reductions observed in blood pressure levels after hydroxyzine administration were not significantly different from those in patients who did not receive pharmacotherapy, indicating that its impact on blood pressure might be attributed to its anxiolytic properties rather than a direct antihypertensive effect | Hydroxyzine works by reducing anxiety, which can indirectly contribute to lowering blood pressure. Anxiety can trigger sympathetic discharge, leading to increased blood pressure. By alleviating anxiety, hydroxyzine may help reduce this sympathetic response. However, the drug's use must be carefully considered, particularly in elderly patients due to potential side effects such as confusion, falls, and other anticholinergic effects |
| **Diazepam<sup>12</sup>** | Thirty-six patients (28 women and 8 men), with excessive hypertension (>190/100 mm Hg), were randomized to receive either oral diazepam, 5 mg (*n* = 17, study group) or sublingual captopril, 25 mg (*n* = 19, control group). Both | Diazepam exerts its antihypertensive effects primarily through its potent anxiolytic properties. As a benzodiazepine, diazepam enhances the effect of the neurotransmitter gamma-aminobutyric acid (GABA) at the GABA-A receptor, |
|  | treatments decreased BP significantly. | resulting in sedative, muscle-relaxant, and anxiolytic effects. This mechanism reduces sympathetic nervous system activity, which is often elevated during periods of high anxiety, thereby lowering blood pressure. Additionally, diazepam's ability to calm patients can mitigate stress-induced hypertensive spikes, providing a dual benefit in managing both anxiety and hypertension |
| **Alprazolam**<sup>21</sup> | 53 patients were enrolled, of whom 27 patients received captopril and 26 patients received alprazolam, alprazolam in a dose of 0.5 mg was as effective as captopril in a dose 25 mg in reducing blood pressure. | Most patients had a high level of anxiety. Anxiety is known to trigger sympathetic discharge, which can lead to increased blood pressure. Alprazolam belongs to the group of benzodiazepines that exhibit anxiolytic, sleep-inducing, sedative, myorelaxant and anticonvulsant effects. |
| **Midazolam**<sup>13</sup> | Midazolam has been shown to significantly reduce blood pressure in acute settings. Studies indicate that midazolam, when administered for | The primary mechanism through which midazolam reduces blood pressure is its potent anxiolytic and sedative effects, which alleviate stress-induced |
|  | hypertensive emergencies, produces substantial reductions in systolic, diastolic, and mean blood pressure. Specifically, midazolam can decrease systolic blood pressure by approximately 23.5%, diastolic by 17.4%, and mean blood pressure by 20.5% | hypertension. By mitigating sympathetic nervous system activation, midazolam effectively lowers elevated blood pressure levels rapidly and safely |

### Limitations and recommendations

This study’s retrospective design and the exclusion of patients with incomplete data present limitations. Future research should include larger sample sizes and prospective analyses to validate these findings. Additionally, there should be a focus on developing standardised protocols for the use of captopril and other antihypertensive agents in emergency settings, particularly for elderly patients and those with comorbid conditions.

While captopril remains a valuable tool for managing acute hypertension in emergency medical settings, its use must be carefully considered, especially in older adults. Further research is warranted to optimise treatment protocols and ensure the safety and efficacy of hypertension management in emergency care.

## Conclusion

Captopril remains a cornerstone treatment for acute hypertension, demonstrating significant efficacy in blood pressure reduction across diverse patient demographics. However, the study highlights the potential role of hydroxyzine, particularly in patients whose hypertension may be exacerbated by anxiety, suggesting that its sedative and anxiolytic properties could offer therapeutic benefits. Despite these observations, the lack of significant statistical evidence and existing medical guidelines cautioning against hydroxyzine use in elderly patients indicate a need for further research. These results advocate for a balanced and individualised approach to hypertensive crisis management, incorporating both established and novel therapeutic strategies to optimise patient outcomes. Although the literature on the topic is limited, we acknowledge the fact of various uses in clinical practice as anxiety is common in case of high blood pressure and may worsen the problem.

Analysis of this data indicates the need for further research into the effectiveness and safety of the drugs used in emergency medical services interventions, especially considering the recommendations for drug use in the elderly and differences in therapeutic approach depending on the type of emergency medical services intervention and reason for the call.

## Data Availability

Not available currently.

## Acknowledgments

None.

## Bibliography

1. Główny Urząd Statystyczny / Obszary tematyczne / Zdrowie / Zdrowie / Pomoc doraźna i ratownictwo medyczne w 2022 roku. (n.d.). Retrieved September 9, 2024, from https://stat.gov.pl/obszary-tematyczne/zdrowie/zdrowie/pomoc-dorazna-i-ratownictwo-medyczne-w-2022-roku,14,7.html.

2. Bloom, A. S. & Schranz, C. Angiotensin-Converting Enzyme Inhibitor–Induced Angioedema of the Small Bowel—A Surgical Abdomen Mimic. J. Emerg. Med. 48, e127–e129 (2015).

3. Gilyarevsky, S. R. et al. Evidence-Based Information Which Could Influence Arterial Hypertension Treatment Approach after Publication of SPRINT Trial Results. Kardiologiia 60, 130–140 (2020).

4. Whelton, P. K. et al. 2017 ACC/AHA/AAPA/ABC/ACPM/AGS/APhA/ASH/ASPC/NMA/PCNA Guideline for the Prevention, Detection, Evaluation, and Management of High Blood Pressure in Adults: A Report of the American College of Cardiology/American Heart Association Task Force on Clinical Practice Guidelines. Hypertension 71, (2018).

5. Van Den Born, B.-J. H., et al. ESC Council on hypertension position document on the management of hypertensive emergencies. Eur. Heart J. - Cardiovasc. Pharmacother. 5, 37–46 (2019).

6. McEvoy, J. W. et al. 2024 ESC Guidelines for the management of elevated blood pressure and hypertension. Eur. Heart J. ehae178 (2024) doi:10.1093/eurheartj/ehae178.

7. Karakiliç, E., Büyükcam, F., Kocalar, G., Gedik, S. & Atalar, E. Same effect of sublingual and oral captopril in hypertensive crisis. Eur. Rev. Med. Pharmacol. Sci. 16, 1642–1645 (2012).

8. Brown, N. J. & Vaughan, D. E. Angiotensin-Converting Enzyme Inhibitors. Circulation 97, 1411–1420 (1998).

9. Schoolwerth, A. C., Sica, D. A., Ballermann, B. J. & Wilcox, C. S. Renal Considerations in Angiotensin Converting Enzyme Inhibitor Therapy: A Statement for Healthcare Professionals From the Council on the Kidney in Cardiovascular Disease and the Council for High Blood Pressure Research of the American Heart Association. Circulation 104, 1985–1991 (2001).

10. Romankiewicz, J. A., Brogden, R. N., Heel, R. C., Speight, T. M. & Avery, G. S. Captopril: An Update Review of its Pharmacological Properties and Therapeutic Efficacy in Congestive Heart Failure. Drugs 25, 6–40 (1983).

11. Leier, C. V. et al. Captopril in primary pulmonary hypertension. Circulation 67, 155–161 (1983).

12. Grossman, E. et al. Antianxiety Treatment in Patients With Excessive Hypertension. Am. J. Hypertens. 18, 1174–1177 (2005).

13. Khorsand, M. R., Enayatrad, M., Yekesadat, S. M., Khodayar, M. & Noyani, A. Comparison of midazolam versus captopril in patients with uncomplicated hypertensive urgency in emergency ward: Double-blind randomized clinical trial. ARYA Atheroscler. 18, 1–8 (2022).

14. Lin, Y.-M. et al. Small Molecule Compound Nerolidol attenuates Hypertension induced hypertrophy in spontaneously hypertensive rats through modulation of Mel-18-IGF-IIR signalling. Phytomedicine 84, 153450 (2021).

15. on behalf of the Swiss Society of Intensive Care Medicine et al. Gender differences in the provision of intensive care: a Bayesian approach. Intensive Care Med. 47, 577–587 (2021).

16. Schröder, W. et al. Gender differences in antibiotic prescribing in the community: a systematic review and meta-analysis. J. Antimicrob. Chemother. 71, 1800–1806 (2016).

17. Tandeter, H. Hypothesis: A single dose of an anxiolitic may prevent unnecessary visits to the emergency room during blood pressure elevations. Med. Hypotheses 88, 35–37 (2016).

18. Philip, R. et al. Variation in hypertension clinical practice guidelines: a global comparison. BMC Med. 19, 117 (2021).

19. By the 2023 American Geriatrics Society Beers Criteria® Update Expert Panel. American Geriatrics Society 2023 updated AGS Beers Criteria® for potentially inappropriate medication use in older adults. J. Am. Geriatr. Soc. 71, 2052–2081 (2023).

20. Williams, B. et al. 2018 ESC/ESH Guidelines for the management of arterial hypertension. Eur. Heart J. 39, 3021–3104 (2018).

21. Yilmaz, S., Pekdemir, M., Tural, Ü. & Uygun, M. Comparison of alprazolam versus captopril in high blood pressure: A randomized controlled trial. Blood Press. 20, 239–243 (2011).

